# Detecting Scoliosis at Scale Using Automated Cobb Angle Analysis in the UK Biobank

**DOI:** 10.1101/2025.03.28.25324832

**Authors:** Marili Niglas, Brandon Whitcher, Dimitri Amiras, Marjola Thanaj, Nicolas Basty, Camilo Bell-Bradford, E Louise Thomas, Jimmy D Bell

## Abstract

**Purpose:** Adult degenerative scoliosis arises after skeletal maturity in an initially normal spine, primarily driven by age-related degeneration. The Cobb angle, the angle between the most tilted vertebrae typically derived from radiographs, remains the clinical standard for assessing curvature severity, yet large-scale evaluation using MRI has not been feasible. This study developed an automated method for Cobb angle estimation from chemical-shift-based water-fat separation (Dixon) MRI and applied it to the UK Biobank to characterise the prevalence and curvature within the general population.

**Methods:** Abdominal Dixon MRI data from 33,889 UK Biobank participants were analysed. Vertebral bone marrow compartments were segmented using a neural network based model, and spinal curvature was quantified using a centroid-based spline-fitting algorithm. Sex-stratified linear regression analyses were performed to explore associations between spinal curvature and anthropometric, socioeconomic, and health-related traits, including back pain and body composition.

**Results:** While scoliosis was clinically diagnosed or self-reported in only 0.5% of participants, the automated approach detected scoliosis (Cobb angle > 10°) in 28%, of which 95% were mild (<25°). Females exhibited higher average Cobb angles and greater curvature across all age groups. Linear regression revealed significant associations between Cobb angle and age, paraspinal muscle fat infiltration, chronic back pain, and visceral adipose tissue in both sexes, and with iliopsoas muscle volume in males only.

**Conclusion:** This fully automated approach enables large-scale, population-based assessment of spinal curvature, revealing adult scoliosis to be substantially under-recognised and closely linked to muscle quality and back pain.

## Introduction

Scoliosis is characterised by an abnormal lateral curvature of the spine. Idiopathic scoliosis, most often diagnosed during adolescence, is the predominant form, whereas in adults it manifests either as residual adolescent idiopathic scoliosis or as de novo degenerative scoliosis following skeletal maturity.

Adult degenerative scoliosis develops after skeletal maturity through asymmetric degeneration of intervertebral discs and facet joints, with extracellular matrix breakdown and inflammation accelerating tissue damage [1,2]. Osteoporosis, sarcopenia, and vertebral fractures further weaken spinal support, contributing to instability [3]. Increasing longevity is driving the prevalence of degenerative scoliosis and demand for treatment [4].

Reported adult degenerative scoliosis prevalence ranges from 2% to 68%, with a pooled prevalence of 38% [5]. The condition is more common in women and adults over 60, although many remain asymptomatic. Symptomatic cases can present with chronic back pain and neurogenic claudication [2]. While radiographic diagnosis of scoliosis could provide more accurate prevalence estimates, large-scale studies to support this analysis were limited until recently.

The diagnostic gold standard for scoliosis is where the Cobb angle - formed between the most tilted vertebral endplates - quantifies curvature severity [6]. Scoliosis is defined radiographically when the Cobb angle exceeds 10° [2]. The severity of spinal deformation guides clinicians in treatment planning and follow-up [7]. Although this technique enables accurate assessment, it requires ionising radiation and manual annotation, leading to inter-observer variability [8]. Automated approaches using vertebral labels [9], corners [10] or spline fitting [8] have been developed to overcome this.

The UK Biobank imaging cohort provides a unique opportunity to assess the prevalence of degenerative scoliosis and explore spinal curvature in relation to conditions and musculoskeletal composition affecting spinal alignment. We applied an automated pipeline to segment the vertebrae bone marrow (VBM) compartments on Dixon MRI and derive Cobb angles using a spline-based centroid method. Furthermore, we investigated the influence of demographic, anthropometric, and musculoskeletal correlates of spinal curvature. This study is intended for epidemiological characterisation, not clinical diagnosis, of scoliosis.

## Methods

### Data

The UK Biobank recruited over 500,000 participants aged 40-85 years between 2006 and 2014 [11]. A subset of 100,000 participants underwent MRI of the heart, brain, and abdomen. The abdominal MRI data were available from 40,755 participants at the time of analysis. The study was approved by the North West Multi-centre Research Ethics Committee (REC reference: 11/NW/0382) with informed consent from all participants. Data access was granted under UK Biobank Application number 44584.

### Image processing and MRI measurements

The abdominal neck-to-knee chemical-shift-based water-fat separation (Dixon) MRI data were processed as previously described [12,13]. Eighteen VBM compartments, spanning the first thoracic to first sacral vertebrae, were manually annotated in 120 participants, and split 80/20 into training/testing subsets. Full segmentation model details are provided in the Supplementary material. The model achieved a Dice similarity coefficient of 0.83 on the test set.

A cubic spline was fitted through the centroids of the VBM compartments using their x-z coordinates to estimate spinal curvature in the coronal plane. Model complexity was set to (n + 1) / 2, where *n* represents the number of VBM compartments. Inflection points from the second derivative of the spline were used to calculate Cobb angles between perpendicular lines to the spline at adjacent points. To capture boundary curvature, additional Cobb angles were measured between terminal vertebrae and their nearest inflection points. The maximum Cobb angle was recorded as the image-derived phenotype (IDP) representing scoliosis severity (Fig. 1).

**Fig. 1.**
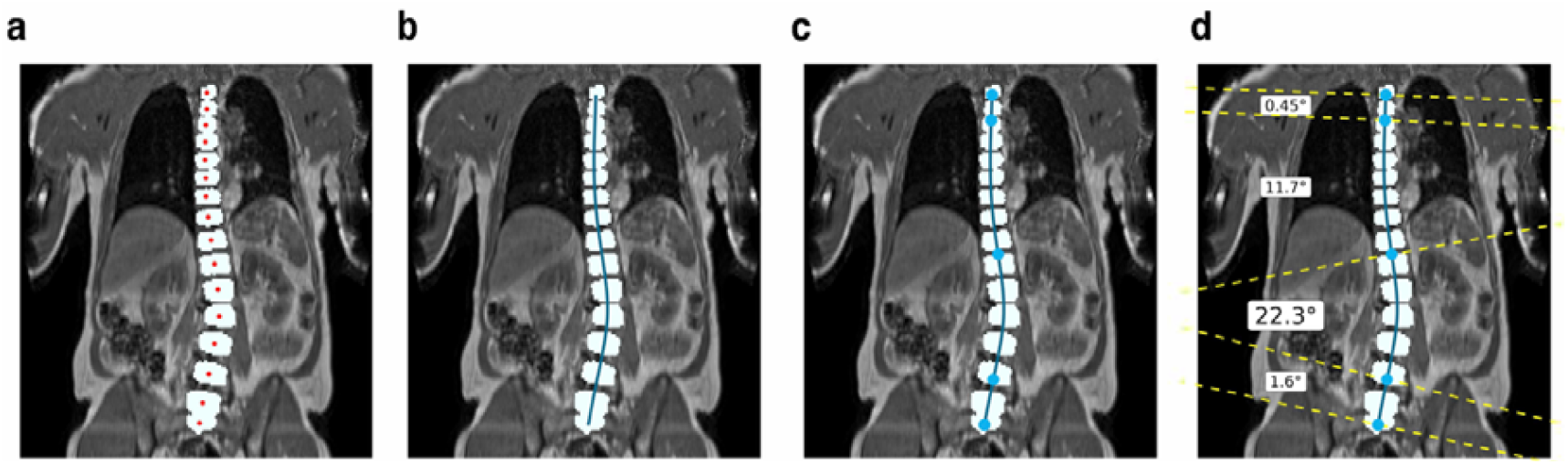
Cobb angle measurement workflow. a) Coronal mid-slice of abdominal MRI of one participant with vertebral bone marrow labels (white) and the corresponding centroids for each compartment (red dots). b) Spline fit through the centroids (dark blue line). c) Points of inflection derived from the second-order derivatives of the fitted spline (light blue dots). d) Cobb angles were estimated by calculating the angle between perpendicular lines to the spline at each pair of adjacent inflexion points throughout the whole spine, where the maximal Cobb angle was taken as a single measure for each participant.

To evaluate relationships between spinal curvature and muscle-fat body compartments, additional IDPs from our previous work [13,14] were included: total skeletal, thigh (left and right) and iliopsoas (left and right) muscle volumes; visceral (VAT) and abdominal subcutaneous adipose tissue (ASAT) volumes; and paraspinal proton density fat fraction (PDFF). The paraspinal muscles included the erector spinae and multifidus muscles.

### Quality control

Visual quality control was performed in 50 participants with the most extreme Cobb angles, a random subset of 50 participants, as well as participants with a scoliosis diagnosis but a Cobb angle <10°. Spurious VBM labels were manually corrected (Supplementary Fig. 1).

Intra-rater reliability and agreement between manual and automated Cobb angle measurements were evaluated using Bland-Altman analysis. A radiologist with 21 years of experience (D.A.) performed manual measurements of Cobb angles and repeated them after an average interval of 11-30 days in 35 participants. Agreement between manual and automated estimates was assessed using the same dataset. Bland-Altman analysis quantified bias and limits of agreement.

### Anthropometric factors and disease definitions

Demographic, lifestyle, biometric, and health data were obtained from the imaging visit and linked healthcare records. Except for sex, ethnicity, Townsend deprivation index, and serum biomarkers, variables were taken from the imaging visit. Ethnicity (field 21000) was classified as White or Non-white, with missing values supplemented by genetic ancestry.

Pre-existing diseases were identified from healthcare records, self-reported medical questionnaires, and UK Biobank specific fields, binarised for presence before imaging. Osteoporosis and osteopenia were defined by diagnostic codes and dual-energy X-ray derived bone mineral density (BMD) T-scores (fields 23204-23301; ≤ −2.5 and −2.5 < T-score ≤ −1, respectively) [15].

### Scoliosis and pain definitions

Participants with scoliosis were identified using UK Biobank fields (Supplementary Table 1). Childhood scoliosis was defined by ICD-10 code M41.1 (juvenile or idiopathic) or a diagnosis before age 18 and was excluded from analysis. Self-reported scoliosis was derived from fields 131907 and 20002. Algorithm-derived scoliosis was defined as a maximum Cobb angle >10° [16] and further categorised by severity: none (0–9°), mild (10–24°), moderate (25–39°), or severe (≥40°) [10].

**Table 1.** Study population demographic characteristics, separated by sex. Values are reported as means with standard deviations for continuous variables, and counts with proportion of total population for overall, female and male cohorts for categorical variables. BMD - bone mineral density; MET - metabolically equivalent task; PDFF - proton density fat fraction.

| Variable | Overall<br>n = 33,889<sup>†</sup> | Female<br>n = 16,907<sup>†</sup> | Male<br>n = 16,982<sup>†</sup> |
| --- | --- | --- | --- |
| **Anthropometric** |  |  |  |
| Ethnicity |  |  |  |
| White | 32,908 (97%) | 16,423 (97%) | 16,485 (97%) |
| Non-white | 981 (2.9%) | 484 (2.9%) | 497 (2.9%) |
| Age (years) | $63.85 \pm 7.68$ | $63.04 \pm 7.48$ | $64.64 \pm 7.80$ |
| Townsend deprivation index | $-1.90 \pm 2.72$ | $-1.83 \pm 2.72$ | $-1.96 \pm 2.71$ |
| Height (cm) | $169.56 \pm 9.23$ | $162.91 \pm 6.22$ | $176.18 \pm 6.61$ |
| Weight (kg) | $76.23 \pm 15.07$ | $68.77 \pm 12.98$ | $83.65 \pm 13.22$ |
| Body mass index (kg/m<sup>2</sup>) | $26.42 \pm 4.32$ | $25.91 \pm 4.69$ | $26.93 \pm 3.85$ |
| Waist circumference (cm) | $88.36 \pm 12.62$ | $82.45 \pm 11.70$ | $94.22 \pm 10.60$ |
| Hip circumference (cm) | $100.65 \pm 8.61$ | $100.63 \pm 9.76$ | $100.66 \pm 7.30$ |
| Waist to hip ratio | $0.88 \pm 0.09$ | $0.82 \pm 0.07$ | $0.93 \pm 0.06$ |
| Hand grip strength - dominant (kg) | $31.75 \pm 10.56$ | $24.31 \pm 5.99$ | $39.09 \pm 8.80$ |
| Moderate-to-vigorous MET (hours/week) | $19.43 \pm 13.58$ | $19.17 \pm 13.40$ | $19.68 \pm 13.75$ |
| Smoking status |  |  |  |
| Never | 21,170 (62%) | 11,150 (66%) | 10,020 (59%) |
| Previous | 11,575 (34%) | 5,269 (31%) | 6,306 (37%) |
| Current | 1,144 (3.4%) | 488 (2.9%) | 656 (3.9%) |
| Alcohol status |  |  |  |
| Never | 1,045 (3.1%) | 669 (4.0%) | 376 (2.2%) |
| Current | 31,729 (94%) | 15,668 (93%) | 16,061 (95%) |
| Previous | 1,107 (3.3%) | 568 (3.4%) | 539 (3.2%) |
| Femur neck BMD (T-score) | -0.67 ± 1.05 | -0.72 ± 1.08 | -0.63 ± 1.03 |
| Lumbar BMD (T-score) | -0.08 ± 1.62 | -0.52 ± 1.49 | 0.35 ± 1.62 |
| **Image derived** |  |  |  |
| Abdominal subcutaneous adipose tissue volume (L) | 8.36 ± 4.07 | 9.74 ± 4.36 | 7.00 ± 3.24 |
| Visceral adipose tissue volume (L) | 3.96 ± 2.31 | 2.77 ± 1.56 | 5.15 ± 2.33 |
| Total muscle volume (L) | 18.00 ± 4.63 | 14.10 ± 1.95 | 21.88 ± 2.99 |
| Iliopsoas muscle volume (L) | 0.65 ± 0.17 | 0.51 ± 0.08 | 0.79 ± 0.12 |
| Thigh muscle volume (L) | 8.56 ± 2.24 | 6.72 ± 1.00 | 10.38 ± 1.51 |
| Paraspinal muscle PDFF (%) | 7.27 ± 3.79 | 7.73 ± 3.97 | 6.81 ± 3.54 |
| **Conditions** |  |  |  |
| Back pain levels |  |  |  |
| Never | 27,358 (81%) | 13,755 (81%) | 13,603 (80%) |
| Acute | 2,245 (6.6%) | 958 (5.7%) | 1,287 (7.6%) |
| Chronic | 4,286 (13%) | 2,194 (13%) | 2,092 (12%) |
| Leg pain | 6,333 (19%) | 3,197 (19%) | 3,136 (18%) |
| Type 2 diabetes mellitus | 1,745 (5.1%) | 554 (3.3%) | 1,191 (7.0%) |
| Osteoporosis | 7,923 (23%) | 4,540 (27%) | 3,383 (20%) |
| Osteopenia | 14,420 (43%) | 7,004 (41%) | 7,416 (44%) |
| Degenerative disc disease | 157 (0.5%) | 90 (0.5%) | 67 (0.4%) |
| Diagnosed scoliosis <sup>2</sup> | 170 (0.5%) | 113 (0.7%) | 57 (0.3%) |
| Cobb angle (°) | 8.72 ± 5.21 | 9.02 ± 5.59 | 8.44 ± 4.79 |
| Subjects with Cobb angle above 10° | 9,497 (28%) | 5,086 (30%) | 4,411 (26%) |
| Severity range |  |  |  |
| None | 24,392 (72%) | 11,821 (70%) | 12,571 (74%) |
| Mild | 9,005 (27%) | 4,776 (28%) | 4,229 (25%) |
| Moderate | 397 (1.2%) | 248 (1.5%) | 149 (0.9%) |
| Severe | 95 (0.3%) | 62 (0.4%) | 33 (0.2%) |

<sup>1</sup> Categorical - count (proportion, %); Continuous - mean ± standard deviation
<sup>2</sup> From hospital admission, primary care sources and self-reported data

Participants reported back pain, its duration, and falls at the imaging visit. Individuals reporting back pain duration ≤3 months were classified as acute and >3 months as chronic [17].

### Study design and cohort

From 40,769 participants with Dixon MRI data, subjects with missing demographic, anthropometric or health-related data were excluded. Non-imaging based variables were age, ethnicity, height, Townsend deprivation index, smoking status, moderate-to-vigorous physical activity (according to metabolic equivalent task values), type 2 diabetes, back pain and leg pain levels, osteoporosis, degenerative disc disease. Imaging based variables were VAT, ASAT, iliopsoas muscle volume and paraspinal muscle PDFF. Participants with juvenile or adolescent scoliosis or who were diagnosed before the age of 18 were excluded. The final study population included 33,889 participants (Supplementary Fig. 2).

### Statistical analysis

Analyses were performed in R v4.3.2 [18]. Continuous variables are reported as mean ± standard deviation (SD), and categorical variables as counts and proportions. Spearman correlation assessed relationships between Cobb angles and other variables. Since Cobb angles ranged from 0° to 90° and lacked circularity, standard linear statistical methods were used.

Multivariable linear regression models were fitted to examine associations between Cobb angles and demographic, anthropometric, and muscle-fat IDPs, adjusting for covariates including age, sex, and physical activity. The Cobb angles were log-transformed, and the covariates were standardised (z-scores). Regression coefficients were reported as standardised β values with 95% confidence intervals and p-values. A Bonferroni-corrected statistical significance threshold of 0.00055 accounted for multiple hypothesis tests.

## Results

Abdominal MRI scans from 40,755 participants were processed, all with successfully fitted splines and derived Cobb angles. After excluding 6,845 participants with missing data and 21 with juvenile scoliosis, the final cohort included 33,889 participants (49.9% female; mean age 63.9 ± 7.7 years) (Table 1).

Representative vertebrae masks, fitted splines and Cobb angle measurements for participants without and with scoliosis are shown in Fig. 2. The average Cobb angle was 8.7 ± 5.2° (range 0.9°-86.6°), with 9.0 ± 5.6° in females and 8.4 ± 4.8° in males (p < 0.00055). Overall, 9,497 participants (28%) had a Cobb angle >10°, 53.6% of whom were female (Fig. 3a). Participants with a clinical or self-reported scoliosis diagnosis (n = 170; 0.5%) had Cobb angles between 1.9° and 84.1°. Of them, 49 had a Cobb angle <10° (Fig. 3b); visual quality control confirmed accurate vertebral segmentation. Using the >10° threshold, scoliosis was identified in 9,376 additional participants, 95.4% of them mild (Fig. 3c). Spinal curvature increased with age and was consistently higher in females across all age groups (Fig. 4; p < 0.00055).

**Fig. 2.**
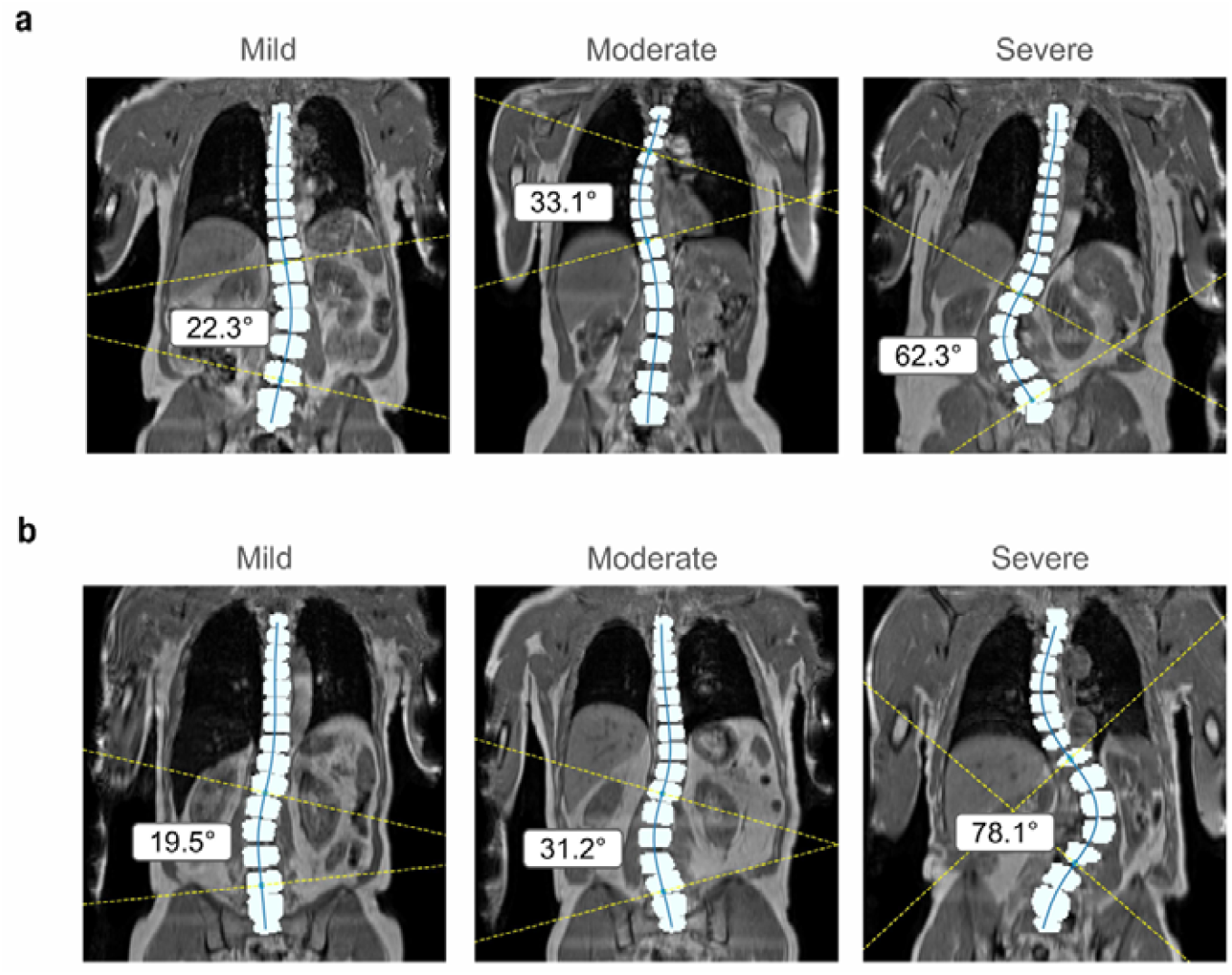
Representative images of participants with a higher than 10-degree maximal Cobb angle with vertebrae masks and fitted spline. a) Participants without a clinical scoliosis diagnosis. b) Participants with a scoliosis diagnosis. Participants were grouped into scoliosis severity categories based on Cobb angle: mild (10-24°), moderate (25-39°), and severe (≥40°).

**Fig. 3.**
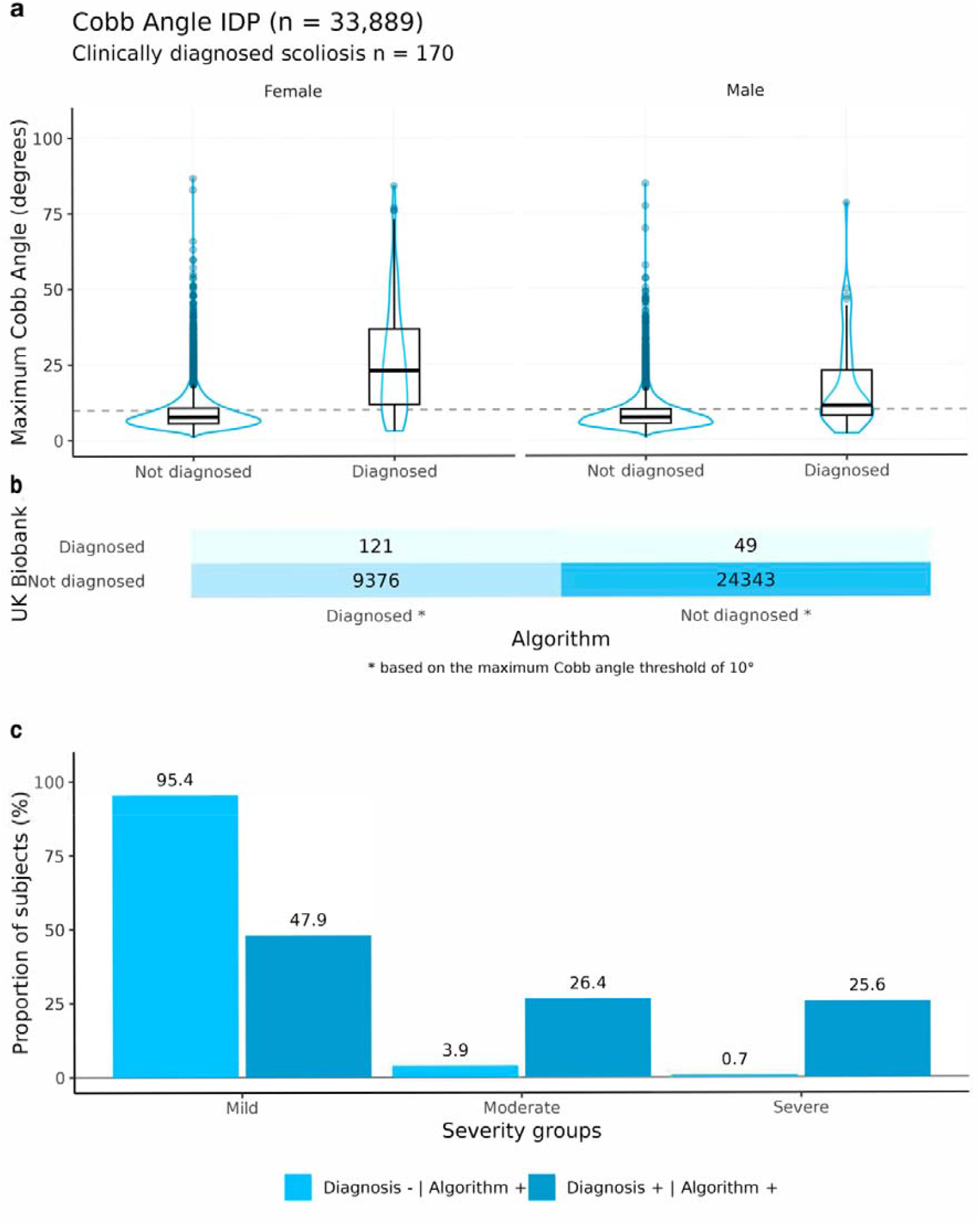
Analysis of Cobb angle by UK Biobank derived clinical scoliosis diagnosis and algorithm-based diagnosis. a) Gender specific distribution of maximal Cobb angle for participants with and without a scoliosis diagnosis code. Algorithm-based scoliosis diagnosis was assigned to participants with a maximal Cobb angle of over 10°, indicated by the dashed line. b) Confusion matrix illustrating agreement between clinical diagnosis and algorithm-based diagnosis for scoliosis. c) Proportion of participants in scoliosis categories according to diagnosis status and scoliosis severity groups. Diagnosis status combined data from hospital records, private healthcare, self-reports, and an algorithm (Cobb angle >10°), distinguishing between cases detected exclusively by the algorithm (Diagnosis - | Algorithm +), cases confirmed by both methods (Diagnosis + | Algorithm +), and cases identified only through healthcare data (Diagnosis + | Algorithm −). Participants were grouped into scoliosis severity categories based on Cobb angle: mild (10-24°), moderate (25-39°), and severe (≥40°).

**Fig. 4.**
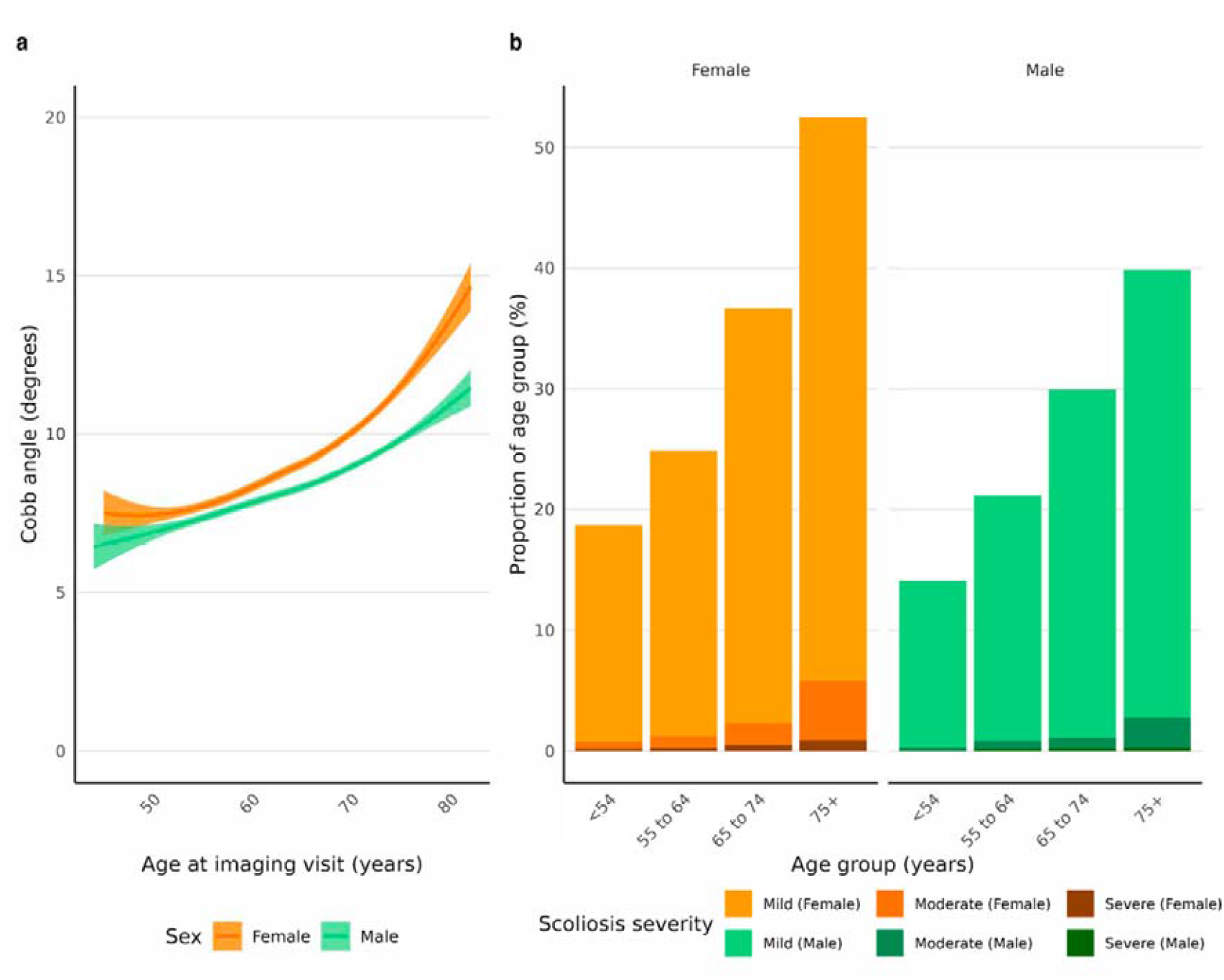
Relationship between Cobb angles to age and sex. a) A line plot with density plots of Cobb angles for age and sex. b) Prevalence of scoliosis by severity group (mild - 10-24°; moderate - 25-39°; severe ≥40°) and age group, stratified by sex.

Spearman correlations with Cobb angle were modest (r < 0.25). Positive correlations were observed for age (r = 0.24 in females; r = 0.21 in males; p < 0.00055) and paraspinal PDFF (r = 0.10 in females; r = 0.07 in males; p < 0.00055). Negative correlations were found for sitting height (r = −0.12 in females; r = −0.10 in males) and iliopsoas muscle volume indexed to height (r = −0.09 in females; r = −0.12 in males; p < 0.00055) (Fig. 5; Supplementary Table 2).

**Table 2.** Cobb angle comparison for female and male participants with back pain. Significance was first tested by one-way ANOVA, and if significant, further post-hoc Tukey test. ^*^ indicate statistically significant for p < 0.05, ^**^ indicate statistically significant after Bonferroni correction.

|  | Female |  |  |  | Male |  |  |  |
| --- | --- | --- | --- | --- | --- | --- | --- | --- |
|  | Never | Acute | Chronic | ANOVA | Never | Acute | Chronic | ANOVA |
| Count (n) | 13,755 | 958 | 2,194 |  | 13,603 | 1,287 | 2,092 |  |
| Cobb angle (°) | 8.9 ± 5.3 | 8.6 ± 4.97 | 10.0 ± 7.4 ** | <0.001 | 8.4 ± 4.7 | 8.2 ± 4.8 | 8.9 ± 5.2 ** | <0.001 |

**Fig. 5.**
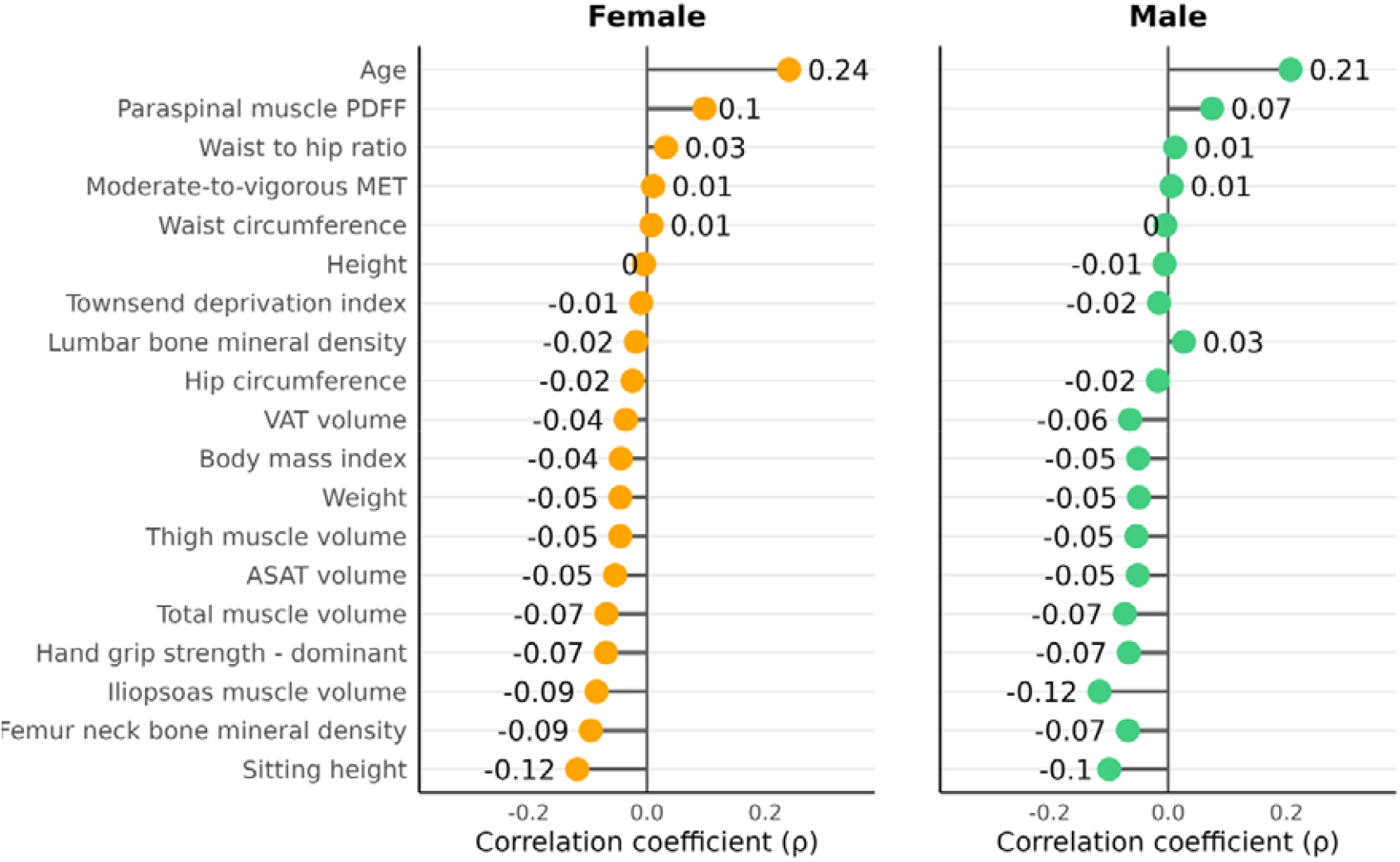
Sex-stratified correlation coefficients between anthropometric variables and Cobb angles. Spearman’s correlation coefficient between the Cobb angle and variables of interest.

Cobb angles were significantly higher in participants reporting chronic back pain compared to those with no or acute pain (p < 0.00055; Table 2). Chronic back pain prevalence rose with scoliosis severity: in females, from 12.2% (no scoliosis) to 40.3 % (severe); in males, from 11.7% to 21.2%. Significant differences were also observed for leg pain (p < 0.00055; Table 3) and osteoporosis (p < 0.00055; Table 4), but not for type 2 diabetes (Supplementary Table 3).

**Table 3.** Cobb angle comparison for female and male participants with leg pain. Significance was first tested by Wilcox rank-sum test and further adjusted by Bonferroni correction. ^*^ indicate statistically significant associations (p < 0.05), ^**^ indicate statistically significant after Bonferroni correction (p < 0.00055).

|  | Female |  | Male |  |
| --- | --- | --- | --- | --- |
|  | No leg pain | Leg pain | No leg pain | Leg pain |
| Count (n) | 13,710 | 3,197 | 13,846 | 3,136 |
| Cobb angle (°) | 8.9 ± 5.5 | 9.5 ± 6.1 ** | 8.4 ± 4.7 | 8.7 ± 5.0 ** |

**Table 4.** Cobb angle comparison for female and male participants with osteoporosis and osteopenia. Significance was first tested by one-way ANOVA, and if significant, further post-hoc Tukey test. ^*^ indicate statistically significant associations (p < 0.05), ^**^ indicate statistically significant after Bonferroni correction (p < 0.00055).

|  | Female |  |  |  | Male |  |  |  |
| --- | --- | --- | --- | --- | --- | --- | --- | --- |
|  | None | Osteopenia | Osteoporosis | ANOVA | None | Osteopenia | Osteoporosis | ANOVA |
| Count (n) | 5,363 | 7,004 | 4,540 |  | 6,183 | 7,416 | 3,383 |  |
| Cobb angle (°) | 8.7 ± 5.2 | 8.8 ± 5.4 | 9.7 ± 6.2 ** | <0.001 | 8.2 ± 4.4 | 8.4 ± 4.7 * | 9.0 ± 5.4 ** | <0.001 |

Sex-stratified regression analysis adjusted for age, height, smoking status, Townsend deprivation index, ethnicity, osteoporosis, degenerative disc disease, pain, and body composition IDPs identified age as the strongest correlate of Cobb angle (p < 0.00055) (Fig. 6, Supplementary Table 4). Chronic back pain (p < 0.00055), paraspinal muscle PDFF (p < 0.00055), and height (p < 0.000549) were also positively associated with spinal curvature. VAT volume (p < 0.00055) and iliopsoas muscle volume (p < 0.00055) in male participants were negatively associated with the Cobb angle. Osteoporosis was positively associated in males only (p < 0.00055). Adjusted R^2^ values (<0.10) indicated that these variables collectively explained <10% of Cobb angle variance.

**Fig. 6.**
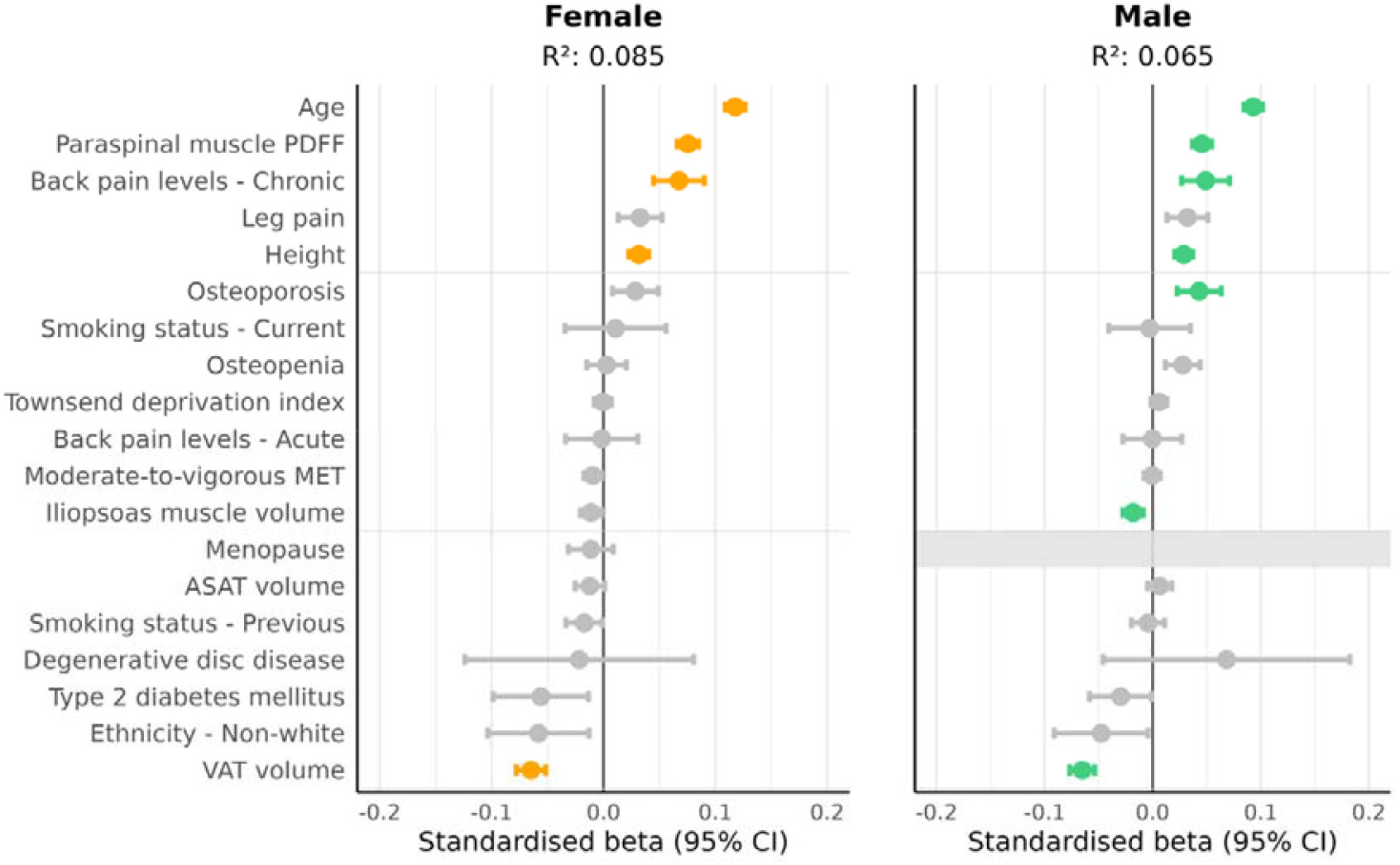
Sex-stratified linear regression models investigating the relationship between Cobb angles and anthropometric variables, image-derived phenotypes and conditions. Linear regression analysis derived standardised regression coefficients (beta) between log-transformed Cobb angles and standardised anthropometric variables, image-derived phenotypes and associated conditions. Colourful beta coefficient labels indicate statistically significant associations (p < 0.000549) after Bonferroni correction.

Automated measurement reliability was evaluated through intra-rater and manual-versus-automated comparisons. The Bland-Altman analysis showed the mean difference between repeated manual measurements (bias = −0.8°, LoA ± 12.66°) was smaller than the difference between manual and automated values (bias = 6.73°, LoA ± 17.0°) (Figure 7).

**Fig. 7.**
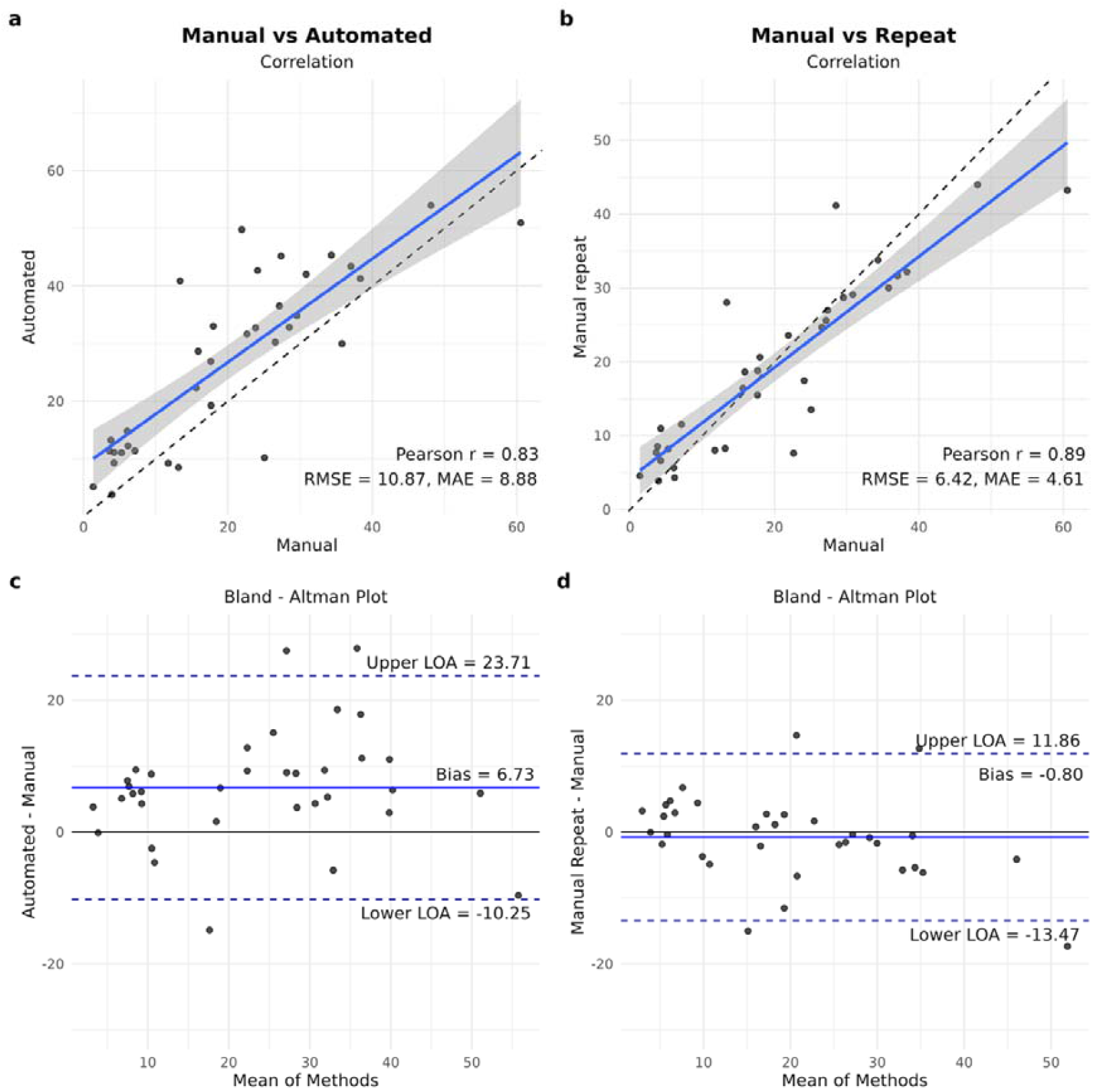
Correlation and agreement analysis between manual and automated measurements. Panels a) and b) show manual against automated value comparisons, and panels c) and d) show initial and repeated manual value comparisons. Panels a) and c) are correlation plots along with corresponding Pearson’s correlation coefficient (r) value, root mean squared error (RMSE) and mean absolute error (MEA). Panels b) and d) are Bland-Altman plots along with the bias line (solid blue) and limits of agreement lines (dashed dark blue).

## Discussion

We developed an automated approach to estimate Cobb angles from Dixon MRI scans, enabling large-scale, radiation-free evaluation of spinal curvature in a population-based cohort. Because Dixon MRI primarily visualises VBM rather than vertebral endplates, we used a centroid-based spline-fitting method to ensure consistent curvature estimation. Using this framework, we quantified the prevalence and severity of scoliosis in a large middle-aged and older UK population and investigated associations with anthropometric traits, muscle and fat IDPs, and musculoskeletal and metabolic conditions.

As part of quality control, small discrepancies between clinical records and algorithm-derived scoliosis assessments were examined. Forty-nine participants had a clinical diagnosis of scoliosis but measured Cobb angle below 10°. Visual inspection confirmed accurate vertebral label segmentation, appropriate spline fitting, and realistic coronal curvature representation. Reproducibility analysis showed marginal overestimation of Cobb angles compared with manual measurements, although the variability remained within typically intra-observer ranges, indicating acceptable agreement for population-level analyses.

Unlike the traditional endplate-based Cobb measurement performed on standing radiographs, this method estimates curvature from centroid trajectories in the supine position. As Cobb angles measured supine are typically 7-12° lower than standing X-rays and no specific threshold has been established for supine imaging, the conventional 10° threshold was retained for comparability. Accordingly, our analyses should be interpreted as epidemiological rather than diagnostic.

We observed a clinically diagnosed scoliosis prevalence of 0.5%, consistent with previous UK Biobank reports (0.8%) [20]. In contrast, the automated MRI-based method identified scoliosis (Cobb angle >10°) in 28% of participants, mostly with mild (10-24°) curvature detectable only through imaging. This discrepancy reflects differences between clinical diagnosis and imaging-derived quantification, highlighting that our approach was designed for population-level assessment rather than clinical diagnosis. Although most cases were mild and likely subclinical, such curvatures may indicate early degenerative spinal changes or age-related biomechanical adaptation.

Female participants exhibited higher Cobb angles and steeper age-related increases than males, consistent with known sex differences in scoliosis prevalence, bone density, muscle mass, and degenerative musculoskeletal conditions [5,21]. After multivariable adjustment, age, paraspinal fat infiltration, chronic back pain, and lower iliopsoas muscle volume remained associated with greater curvature, whereas VAT volume showed an inverse relationship. These findings suggest that both muscular degeneration and body composition influence spinal alignment. However, the regression models explained <10% of Cobb-angle variance, indicating additional influences such as vertebral geometry, posture, and genetic predisposition.

Asymmetrical mechanical loads can induce or exacerbate scoliosis by affecting vertebral bodies, intervertebral discs, and surrounding musculature. Several studies underscore the stabilising role of torso muscles, particularly iliopsoas and paraspinal muscles, with paraspinal fat infiltration linked to higher Cobb angles [19,21,22]. Zhou et. al. reported in 50 adult degenerative scoliosis patients that fat infiltration of all torso stabilisers correlated with Cobb angle in men, whereas only fat infiltration of the paraspinal muscles correlated in women [22]. Consistent with this, we observed positive associations between paraspinal fat infiltration and Cobb angle in both sexes, yet a negative association between iliopsoas muscle volume and curvature in men only. These findings suggest that women may rely more on paraspinal than psoas muscles for coronal spinal stabilisation.

Chronic back pain prevalence increased with scoliosis severity in both sexes and remained significant after adjusting for known risk factors such as age, degenerative conditions, and muscle composition [3,19,23]. While abnormal spinal alignment is recognised as a risk factor for back pain [3,24], our findings underscore the added importance of quantifying spinal curvature severity. The high proportion of participants without a formal scoliosis diagnosis supports using continuous measures such as the Cobb angle rather than diagnostic labels in future studies of back pain and spinal alignment.

Several limitations of this study should be acknowledged. First, MRI was acquired in the supine position, where Cobb angles are typically 7-12° lower than standing radiographs [19], potentially underestimating scoliosis prevalence when applying a 10° threshold. Second, bone mineral density (BMD) was incorporated indirectly via osteoporosis and osteopenia classifications. Despite including key factors such as age, torso muscle volume and quality, and osteoporosis, linear regression models explained <10% of Cobb angle variance. This suggests additional influences including genetic predisposition [25], biomechanical characteristics such as facet joint shape and orientation [23], and postural asymmetries leading to uneven spinal loading [7], may play important roles in scoliosis development and progression. Finally, distinguishing adult-onset from residual adolescent scoliosis remains challenging despite excluding known adolescent diagnoses.

In conclusion, this study presents an automated, radiation-free method to estimate Cobb angles from Dixon MRI, enabling large-scale assessment of spinal curvature. The approach revealed a higher prevalence of mild, previously undiagnosed scoliosis than clinically reported. Associations with age, muscle quality, and back pain highlight the interrelation between spinal alignment and musculoskeletal health. Although limited by supine imaging and low explained variance, this scalable method provides a valuable tool for population studies of spinal ageing and health.

## Acknowledgements

The authors would like to thank Frances Williams and Liliana Szabo for input on the manuscript. We would also like to thank the participants in the UK Biobank imaging study. This research was carried out using the UK Biobank Application number 44584, and funded by a Calico Life Sciences LLC research grant.

## Author contributions

M.N.: Data curation, Formal analysis, Investigation, Methodology, Software, Validation, Visualization, Writing - original draft, Writing - review & editing. B.W.: Conceptualization, Data curation, Formal analysis, Methodology, Software, Supervision, Writing - review & editing. D.A.: Data curation, Validation, Writing - review & editing. M.T.: Data curation, Investigation, Writing - review & editing. N.B.: Conceptualization, Data curation, Investigation, Methodology, Writing - review & editing. C.B.B.: Data curation; Validation, Writing - review & editing. E.L.T.: Conceptualization, Supervision, Writing - review & editing. J.D.B.: Conceptualization, Project administration, Supervision, Writing - review & editing.

## Declarations

Fully anonymised images and participant metadata were obtained through UK Biobank Access Application number 44584. The UK Biobank has approval from the North West Multi-Centre Research Ethics Committee (REC reference: 11/NW/0382), and obtained written informed consent from all participants before the study. All methods were performed in accordance with the relevant guidelines and regulations as presented by the appropriate authorities, including the Declaration of Helsinki.

## Data Availability

Our research was conducted using UK Biobank data. Under the standard UK Biobank data sharing agreement, we (and other researchers) cannot directly share raw data obtained or derived from the UK Biobank. However, under this agreement, all of the data generated, and methodologies used in this paper are returned by us to the UK Biobank, where they will be fully available. Access is obtained directly from the UK Biobank to all bona fide researchers upon submitting a health-related research proposal to the UK Biobank https://www.ukbiobank.ac.uk.

