## Supplementary material for "Detecting Scoliosis at Scale Using Automated Cobb Angle Analysis in the UK Biobank"

Image processing

Organ segmentation

We adapted a recent three-dimensional version of the U-Net architecture [1], originally developed for label-free segmentation in 3D microscopy [2]. All model components were implemented in TensorFlow (TF). Because the available training data were limited, the network was optimised to generate reliable segmentations suitable for quality control from minimal samples. To maximise data efficiency, we adopted a multi-task learning strategy [3]. The network was supervised through binary output heads rather than a multi-class scheme, as several anatomical compartments can spatially overlap. Multiple compartments and organs were annotated in the same individuals, but cross-dataset comparison was avoided, as such results could be influenced by differences in participant selection, annotation conventions, or image acquisition protocols.

The implemented 3D U-Net used 72 feature channels at the outer layers, with a maximum of 1,152 channels at deeper levels. Skip connections used concatenation, and upsampling was performed via transposed convolutions. A customised data-handling pipeline efficiently streamed large volumes of data and performed on-demand augmentation. To reduce computational load, 3D multi-channel images were stored as unrolled PNGs within TFRecords, enabling parallelised random batching using TensorFlow’s built-in tools. Data augmentation occurred dynamically on the GPU during training. The batch size was six, requiring tailored engineering to handle large tensors within GPU memory limits.

Each voxel contained five input channels—fat, water, in-phase, out-of-phase, and a body mask indicating inclusion within the body contour. Separate logit heads were trained for each organ, using a combined loss function of Dice coefficients [4] and binary cross-entropy.

Validation loss curves demonstrated that batch normalisation and on-the-fly augmentation provided adequate regularisation. The model was trained on 80,000 patches of size 96×96×96 voxels, using the Adam optimiser with a learning rate decaying quadratically from 1×10⁻⁵ to 1×10⁻⁷. During inference, voxel-level binary segmentation for each organ was obtained using Otsu thresholding [5].

Data augmentation

Data augmentation incorporated smooth three-dimensional deformations that transformed volumes as coherent 3D objects rather than slice by slice. Small voxel batches were sampled iteratively, assigned random Gaussian values, and the resulting noise was convolved with Gaussian filters of random widths. The combined field was added dynamically to the raw image as a noise vector. Additional augmentation included smooth elastic warping, where each voxel received a continuous 3D optical flow offset in random spatial directions. This approach effectively captured a heterogeneous range of local spatial distortions. The same warping function was simultaneously applied to segmentation masks to maintain spatial alignment between training inputs and supervision labels.

Each voxel value was derived from a location displaced by an optical flow vector sampled from a Gaussian process. Neighbouring voxels shared highly correlated offsets to preserve local structural continuity, while distant voxels had weaker correlations. To limit computational load during optical flow sampling, images were cropped to a 174×174×174 window, within which a 4×4×4 lattice (64 fixed points) was defined. Pairwise distances between lattice points were used to construct a covariance matrix describing the spatial correlation of deformations. A Gaussian kernel with 24-voxel width was applied, and the resulting 3×64 values were scaled randomly (uniformly sampled in [0,4]) to generate optical flow vectors along the three spatial dimensions. These vectors were interpolated across the entire image using a polyharmonic spline, and voxel intensities were resampled at their displaced 3D positions. A central 96×96×96 patch from the warped image was used for training.

Segmentation masks were converted to floating-point probability maps before warping, with clipping heuristics applied post-resampling to maintain valid probability ranges. Final organ or tissue volumes were obtained by thresholding model outputs, removing disconnected regions, and multiplying the remaining voxel counts by image resolution.

Quality control involved repeated visual inspection of extreme predicted volumes for each organ, along with random reviews of hundreds of subjects. The training dataset was iteratively expanded to include problematic examples, and the model retrained until extreme or random cases no longer displayed outlier segmentations. Further architectural details are provided in reference [6].

Supplementary figures

**Supplementary Fig. 1** Representative image of spurious vertebrae label correction. A - Subject abdominal MRI with the original vertebrae labels, where the white arrows indicate spurious vertebrae masks. B - Manually corrected vertebrae labels. Maximal Cobb angle for the subject is shown to the left of the vertebral column along with the tangent lines crossing at the spine spline at inflexion points.

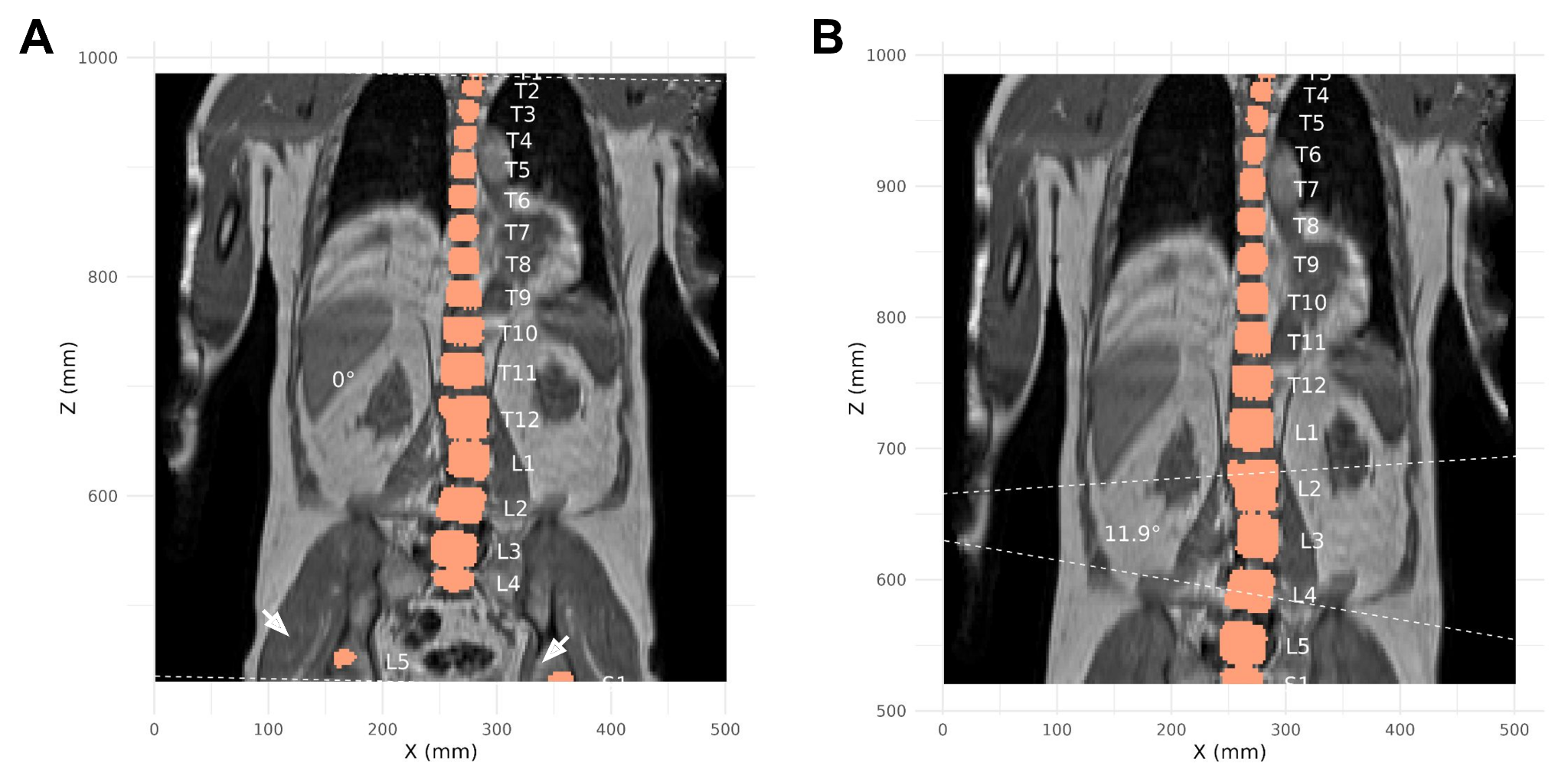

**Supplementary Fig. 2** A flow chart illustrating the selection of participants from the UK Biobank for subsequent analysis.

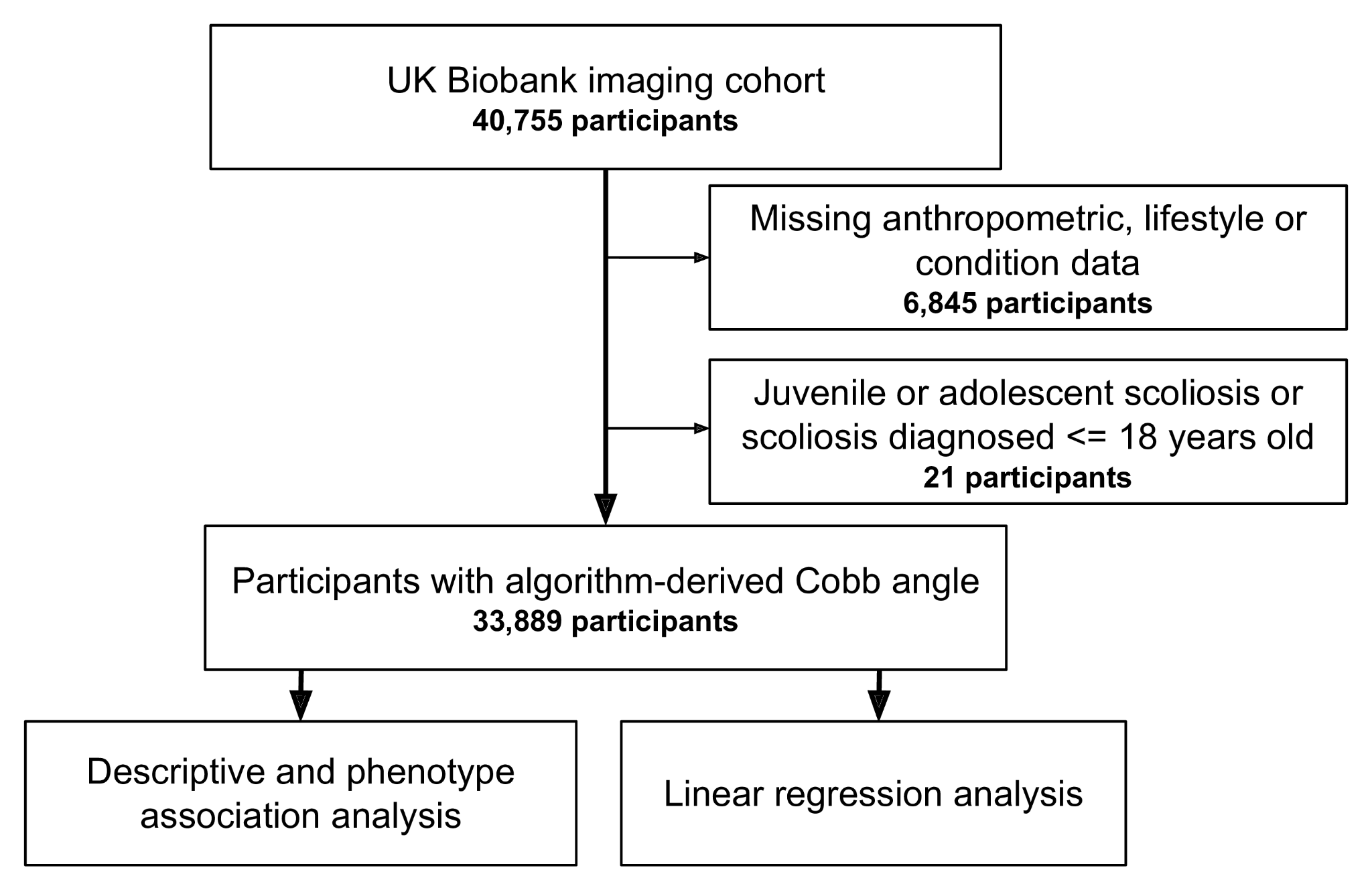

Supplementary tables

**Supplementary Table 1** Diagnostic codes and UK Biobank (UKB) fields used for identifying participants with scoliosis diagnosis. The subjects were identified using the International statistical Classification of Disease 9 and 10 (ICD9, ICD10) summary diagnoses (main and secondary). Primary care, hospital admission and self-reported data were derived from UKB-specific fields of 131907 and 20002.

| **Scoliosis type** | **Scoliosis codes** | **Subjects (n)** | **Proportion of identified (%)** | **Childhood scoliosis (n)** |
| --- | --- | --- | --- | --- |
| Kyphoscoliosis and scoliosis | ICD9: 737.3 | 0 | 0 | 0 |
| Congenital musculoskeletal deformities of the spine | ICD9: 754.2 | 0 | 0 | 0 |
| Congenital scoliosis due to congenital bony malformation | ICD10: Q76.3 | 0 | 0 | 0 |
| Postradiation scoliosis | ICD10: M96.5 | 0 | 0 | 0 |
| Infantile idiopathic scoliosis | ICD10: M41.0 | 0 | 0 | 0 |
| Juvenile and adolescent idiopathic scoliosis | ICD10: M41.1 | 1 | 0.5 | 1 |
| Other idiopathic scoliosis | ICD10: M41.2 | 1 | 0.5 | 0 |
| Thoracogenic scoliosis | ICD10: M41.3 | 2 | 1 | 0 |
| Neuromuscular scoliosis | ICD10: M41.4 | 0 | 0 | 0 |
| Other secondary scoliosis | ICD10: M41.5 | 0 | 0 | 0 |
| Other forms of scoliosis | ICD10: M41.8 | 11 | 5.8 | 2 |
| Scoliosis, unspecified | ICD10: M41.9 | 58 | 30.4 | 2 |
| Unspecified scoliosis | UKB field 131907, hospital and primary care records | 52 | 27.2 | 0 |
| Unspecified scoliosis | UKB field 131907, self-reported | 66 | 34.6 | 16 |
| Self-reported scoliosis | UKB field 20002,  code 1535 | 0 | 0 | 0 |
| Total |  | 191 | 100 | 21 |

**Supplementary Table 2** Spearman correlation coefficients (ρ) between Cobb angle and anthropometric variables for male and female participants. Spearman’s rank correlation values range from -1 (strong negative correlation) to +1 (strong positive correlation). ρ - Spearman's correlation coefficient, rho; p-adj - Bonferroni adjusted p-value. * indicate statistically significant associations (p < 0.05), ** indicate statistically significant after Bonferroni correction (p < 0.00055).

|  | **Female** | **Male** |
| --- | --- | --- |
| **Variable** | **ρ** | **ρ** |
| **Anthropometric** |  |  |
| Townsend deprivation index | -0.010 | -0.015 * |
| Age | 0.240 ** | 0.207 ** |
| Height | -0.005 | -0.006 |
| Sitting height | -0.118 ** | -0.099 ** |
| Weight | -0.045 ** | -0.049 ** |
| Body mass index (kg/m²) | -0.044 ** | -0.050 ** |
| Waist circumference | 0.008 | 0. |
| Hip circumference | -0.024 * | -0.013 * |
| Waist to hip ratio | 0.032 ** | 0.012 |
| Hang grip strength - dominant (kg) | -0.069 ** | -0.066 ** |
| Moderate-to-vigorous MET | 0.010 | 0.006 |
| Femur neck BMD | -0.095 ** | -0.068 ** |
| Lumbar BMD | -0.019 * | 0.027 * |
| **Image derived (MRI)** |  |  |
| Abdominal subcutaneous adipose tissue volume (L) | -0.054 ** | -0.051 ** |
| Visceral adipose tissue volume (L) | -0.036 ** | -0.064 ** |
| Total muscle volume (L) | -0.068 ** | -0.073 ** |
| Iliopsoas muscle volume (L) | -0.085 ** | -0.116 ** |
| Thigh muscle volume (L) | -0.045 ** | -0.054 ** |
| Paraspinal muscle PDFF (%) | 0.097 ** | 0.074 ** |
| ^1^ Mean ± SD; n (%); Count (proportion %) | | |
| *^2^* From hospital admission and primary care sources, and self-reported instances | | |

**Supplementary Table 3** Cobb angle comparison for female and male participants with type two diabetes (T2D). Significance was first tested by Wilcox rank-sum test and further adjusted by Bonferroni correction. * indicate statistically significant associations (p < 0.05), ** indicate statistically significant after Bonferroni correction (p < 0.00055).

|  | **Female** | | **Male** | |
| --- | --- | --- | --- | --- |
|  | **No T2D** | **T2D** | **No T2D** | **T2D** |
| Count (n) | 16,353 | 554 | 15,791 | 1,191 |
| Cobb angle (°) | 9.0 ± 5.6 | 8.6 ± 5.0 | 8.5 ± 4.8 | 8.1 ± 4.1 |

**Supplementary Table 4** Standardised beta coefficients from sex-stratified linear regression models. Data is presented as standardised beta coefficient (St. beta) and 95% confidence intervals (CI). Smoking status, alcohol intake and back pain levels were compared to reference group ‘Never’, ethnicity was compared to reference group ‘White’, conditions such as degenerative disc disease, osteopenia, osteoporosis, leg pain, type 2 diabetes mellitus, and menopause were compared to reference group ‘None’. Last two rows indicate the R-squared and adjusted R-squared values of each of the linear regression models. * indicate statistically significant associations (p < 0.05), ** indicate statistically significant after Bonferroni correction (p < 0.00055).

|  | **Female** n = 16,907 | | **Male** n = 16,982 | |
| --- | --- | --- | --- | --- |
| **Characteristic** | **St. beta** | **95% CI** | **St. beta** | **95% CI** |
| Ethnicity | -0.06 * | -0.10, -0.01 | -0.05 * | -0.09, 0.00 |
| Age | 0.12 ** | 0.11, 0.13 | 0.09 ** | 0.08, 0.10 |
| Townsend deprivation index | 0.00 | -0.01, 0.01 | 0.01 | 0.00, 0.01 |
| Height | 0.03 ** | 0.02, 0.04 | 0.03 ** | 0.02, 0.04 |
| Moderate-to-vigorous MET | -0.01 * | -0.02, 0.00 | 0.00 | -0.01, 0.01 |
| Smoking status |  |  |  |  |
| Previous | -0.02 * | -0.03, 0.00 | 0.00 | -0.02, 0.01 |
| Current | 0.01 | -0.03, 0.06 | 0.00 | -0.04, 0.03 |
| Paraspinal muscle PDFF | 0.08 ** | 0.07, 0.09 | 0.05 ** | 0.04, 0.05 |
| Iliopsoas muscle volume | -0.01 * | -0.02, 0.00 | -0.02 ** | -0.03, -0.01 |
| Visceral adipose tissue volume | -0.06 ** | -0.08, -0.05 | -0.07 ** | -0.08, -0.05 |
| Abdominal subcutaneous adipose tissue volume | -0.01 | -0.03, 0.00 | 0.01 | -0.00, 0.02 |
| Back pain levels |  |  |  |  |
| Acute | 0.00 | -0.03, 0.03 | 0.00 | -0.03, 0.03 |
| Chronic | 0.07 ** | 0.04, 0.09 | 0.05 ** | 0.03, 0.07 |
| Leg pain | 0.03 * | 0.01, 0.05 | 0.03 * | 0.01, 0.05 |
| Osteopenia | 0.00 | -0.01, 0.02 | 0.03 * | 0.01, 0.04 |
| Osteoporosis | 0.03 * | 0.01, 0.05 | 0.04 ** | 0.02, 0.06 |
| Degenerative disc disease | -0.02 | -0.12, 0.08 | 0.07 | -0.05, 0.18 |
| Type 2 diabetes mellitus | -0.06 * | -0.10, 0.01 | -0.03 * | -0.06, 0.00 |
| Menopause | -0.01 | -0.03, 0.01 | - | - |
| R² | 0.085 | | 0.065 | |
| Adjusted R² | 0.084 | | 0.064 | |
